# aiDIVA – Diagnostics of Rare Genetic Diseases Using Large Language Models

**DOI:** 10.1101/2025.09.04.25335099

**Authors:** Dominic Boceck, Lucia Laugwitz, Marc Sturm, Daniela Bezdan, Axel Gschwind, Tobias B. Haack, Stephan Ossowski

## Abstract

Genome sequencing (GS) enables the accurate identification of genetic variants in most genomic regions and is rapidly transforming routine diagnostics for rare diseases (RD). While streamlined data generation is scalable, efficient prioritization and correct clinical interpretation of detected alterations remain a challenge, often requiring manual classification by experts with years of training. Hence, there is a need for AI-driven clinical decision support systems that assist clinical experts in identifying causal variants or, in case of large-scale re-analysis of unsolved cases, fully automate the process. To this end, many tools have been developed to estimate the impact of variants on protein function. However, only a small number of tools combine genomic data, variant annotations, and phenotypic data to diagnose cases.

Here we introduce aiDIVA, an ensemble-AI featuring a hierarchically organized set of statistical and machine learning models trained on genomic and phenotypic data to identify the causal variant(s) among tens of thousands of genetic variants of a patient. aiDIVA generates pathogenicity classifications for each variant using a random forest AI model and an evidence-based score for dominant and recessive diseases. It combines these predictions with additional clinical metadata to prioritize and rank the most likely causal variants. aiDIVA uses large language models (LLMs) to further improve and explain the results. Finally, the aiDIVA-meta model combines all scores to generate a ranked list of variants. In a benchmark analysis on more than 3,000 diagnostically solved RD patients, the causal variant was included in 97% of the cases among the top-3 candidate variants reported by aiDIVA-meta. Unlike comparative methods, aiDIVA provides interpretable explanations for the best candidates.

## Introduction

Advances in sequencing technologies made genome sequencing (GS) feasible for clinical research and diagnostics. However, the unbiased detection of millions of germline variants per affected individual necessitates sophisticated prioritization strategies for the clinical interpretation of identified variants^1^. Despite recent improvements, the diagnostic rate for most rare genetic diseases remains below 40%, and numerous patients and their families continue their diagnostic odyssey^2,3^. Moreover, the growing complexity of genomic datasets and their analysis options may even exacerbate the bottleneck in final clinical evaluation and prolong the time to diagnosis and personalized treatment. Depending on the data background and clinical context, hundreds of variants may require manual evaluation despite rigorous bioinformatic pre-processing and filtering^4^. Artificial intelligence (AI) is widely believed a potential avenue to address the ever-increasing complexity and numbers of diagnostic requests with limited human resources. Moreover, AI has the potential to fully automate the recommended reanalysis of existing genetic datasets of initially undiagnosed cases at scale^3^. Ideally, such AI methods integrate standardized genotype and phenotype information and suggest a small list of candidate variants per individual for subsequent validation by clinical experts.

Over the last decade, guidelines have been developed to assess the pathogenicity of genetic variants. The commonly used American College of Medical Genetics and Genomics (ACMG) guidelines^5^ categorize variants into five tiers: pathogenic, likely pathogenic, uncertain significance (VUS), likely benign, and benign. The classification process involves evaluating various types of evidence, including gene-level data, population data, computational predictions, functional data, and segregation information. Several tools that semi-automate the evaluation of ACMG criteria for specific subsets of disorders and genes are already available, however with a highly variable diagnostic accuracy^6,7^. Furthermore, machine learning (ML) has been shown to improve pathogenicity prediction (ACMG criterion PP3) for small coding variants (e.g. PolyPhen^8,9^, FATHMM-XF^10^, CAPICE^11^, and REVEL^12^). These models are typically trained on variants with established ACMG classifications, e.g., from ClinVar^13^ or HGMD^14^. Usually, these models include a wide range of features, such as population allele frequency, evolutionary conservation or structural effects on RNAs and proteins. However, studies have shown that model training is often biased due to data circularity when repeatedly using the same training and evaluation data for different models^15^. To reduce this bias, the Eigen classifier used an unsupervised model trained on unlabeled data^16^, whereas CADD used a logistic regression model trained on simulated *de novo* variants^17,18^. AlphaMissense^19^, a recent addition to the portfolio, combines protein structure predictions from AlphaFold^20^ with population frequency data and other genomic features to improve variant classification.

ML methods require several precomputed features generated by other tools or models, which are often incomplete (many pathogenicity predictors are only computed for coding variants) or outdated (some protein damage predictors are based on HG19 and must be lifted to newer assemblies). Extracting or updating features based on other functional impact models can therefore be tedious. Another shortcoming of most functional impact predictors is their focus on single nucleotide variants (SNVs), whereas the classification of small in-frame insertions and deletions (indels) are mostly not included (e.g. AlphaMissense^19^, REVEL^12^). Only a few indel-specific algorithms are available, such as CADD-Indel^17,18^, CAPICE-Indel^11^, and FATHMM-indel^21^. In clinical practice, most indel-specific tools do not perform sufficiently well to be included in a diagnostic workflow. Additionally, specialized tools, such as SpliceAI^22^, are required for the classification of splicing variants and only recently an AI model, PromoterAI, was published specifically targeting variants located in promoters^23^.

Despite continuous advancements in *in silico* prediction tools, the correct assignment of causal variants remains reliant on the correlation with detailed phenotypic information (ACMG criterion PP4). Tools that integrate phenotypic and molecular characteristics to pinpoint the causal variants underlying a given rare Mendelian disease are scarce. Exomiser^24^ is a frequently used tool that applies logistic regression to combine genomic and evolutionary features with observed phenotypes described according to the Human Phenotype Ontology (HPO)^25^. Exomiser ranks variants according to their likelihood of being causal for a disorder and an automated ACMG classification has been added to the latest version^26^. Other methods, such as eDiVA^27^ or AI-MARRVEL^28^, use a random forest (RF) approach to generate a pathogenicity score based on multiple genomic and evolutionary features before combining them with HPO terms and segregation information in separate steps. DeepPVP^29^ employs a deep neural network to incorporate genomic, evolutionary, phenotypic, and inheritance information to compute a binary label (“causative” or “non-causative”) for each variant.

Recent improvements in the field of generative AI and the advent of reliable large language models (LLM) have led to increased efforts in developing AI-assisted diagnostics methods^30^. Training and fine-tuning of such models are facilitated by the vast amount of information found in clinical variant databases, ontologies, and publications. Recent studies have investigated the potential of LLMs to prioritize causal genes based only on phenotypic information. Kim et al.^31^ asked pre-trained LLMs for a list of the most suitable candidate genes responsible for a disease phenotype described in the prompt. However, this approach only returned the causal gene among the top 50 candidates in 17.0 % of the test cases, leaving room for improvement. The Articulate Medical Intelligence Explorer (AMIE)^32^, is an LLM using clinical data (clinical history, physical examination, investigations and procedures) to generate a differential diagnosis with good accuracy (top-10 accuracy 59.1%).

Here, we introduce aiDIVA, a tool that combines statistics- and ML-based pathogenicity classifiers (evidence-based and predictive models) with pre-trained LLMs, such as GPT-4o^33^, Llama^34^, or Mistral^35^. This ensemble-AI approach integrates genomic, evolutionary, clinical, phenotypic, and inheritance information to rank the most likely causal variants and provides reasoning for suggested variants. ML models were trained and tested using more than 150,000 known benign or pathogenic variants, as well as inhouse cases diagnosed by clinical experts. Moreover, aiDIVA utilizes a specialized model for in-frame indels.

We assessed aiDIVA using a benchmark set of more than 3,000 RD cases curated in accredited routine diagnostics. aiDIVA identified the causal variants among the top-3 ranked candidates in 97.0 % of the cases. Furthermore, we performed a comprehensive benchmark comparison with Exomiser and AI-MARRVEL.

## Results

### Pathogenicity Classification and Ranking of Most-Likely Causal Variants using aiDIVA

We present benchmarking results for aiDIVA, a newly developed AI platform, that integrates statistical models, ML classifiers, and LLMs to classify variant pathogenicity and prioritize causal variants in RD cases (**Figure 1**). aiDIVA comprises two complementary models for pathogenicity scoring: The first component is an evidence-based model which leverages curated information from the scientific literature, established disease-associated variant databases such as ClinVar, HGMD, and OMIM and gene-phenotype matching (**Table 1**). This model is loosely based on the ACMG guidelines and supports separate statistical frameworks for dominant and recessive Mendelian inheritance, implemented as *aiDIVA-EB-dominant* and *aiDIVA-EB-recessive*, respectively. The second component (aiDIVA-RF, **Figure 1**, **Supplemental Figure 1**) is based on a RF classifier that integrates a broad range of genomic and clinical biomarkers. This model not only performs pathogenicity classification but also incorporates phenotype-driven, disease-similarity-based ranking to identify the most likely causal variants.

**Figure 1.**
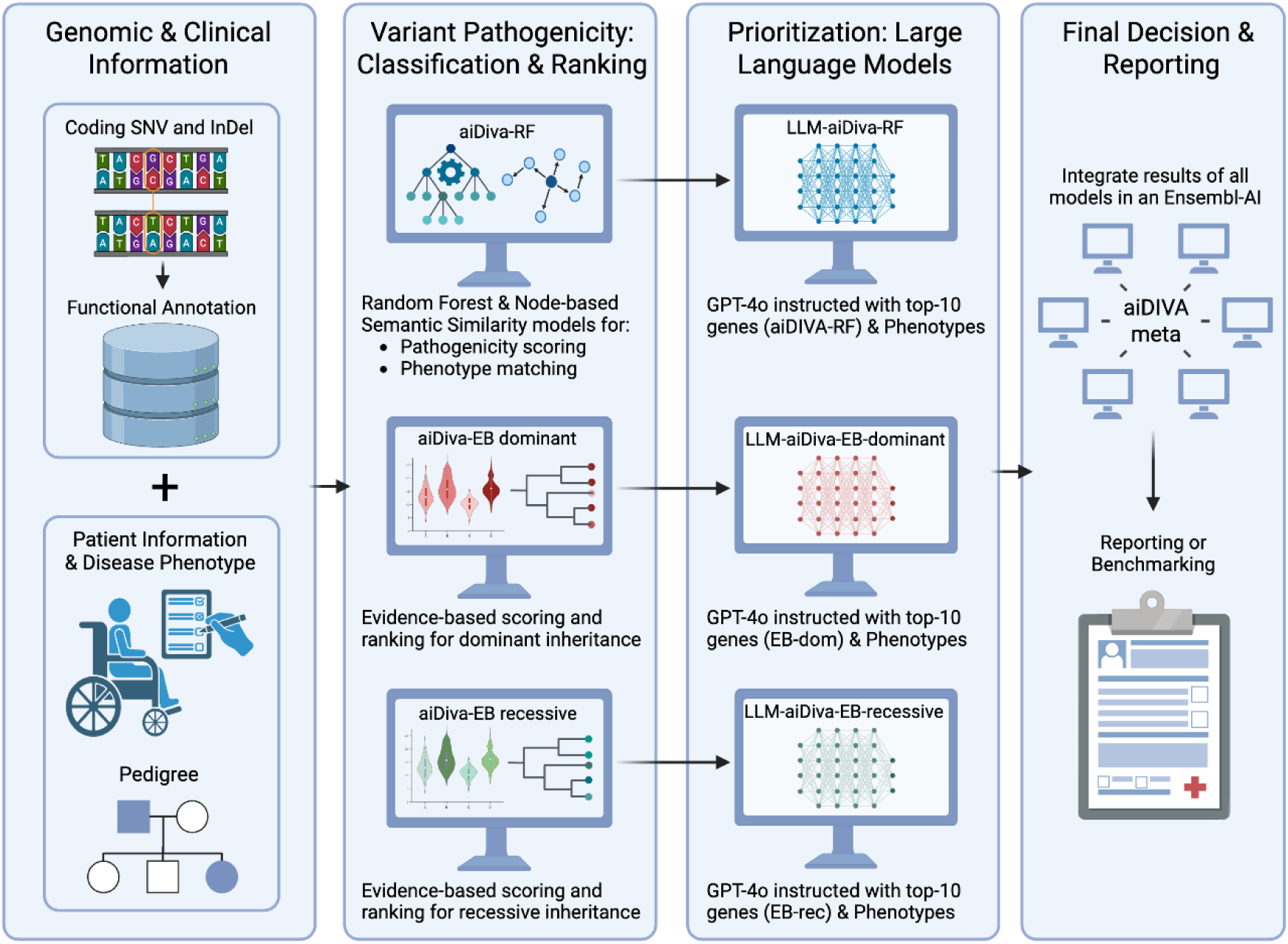
Workflow of the aiDIVA pipeline. First, three different variant rankings are created (aiDIVA-RF, aiDIVA-EB-dominant, and aiDIVA-EB-recessive). These different rankings are used as basis for a refinement with LLMs (LLM-aiDIVA-RF, LLM-aiDIVA-EB-dominant, and LLM-aiDIVA-EB-recessive). Finally, the rankings and refinements are combined using a random forest model (aiDIVA-meta), resulting in a final ranking of causal variant candidates reported to the clinical expert. *Created in BioRender. Ossowski, S. (2025)* https://BioRender.com/wc5hp11

**Table 1.** Scoring criteria for the dominant and recessive models. Most criteria or points are identical for the recessive and the dominant models. Only criteria marked with an asterisk differ between the models.

| Criterion | points (dominant) | points (recessive) |
| --- | --- | --- |
| HPO term matches between patient and variant gene | $1.0 + \sqrt{(\# \text{matches})}$ if there are matches | $1.0 + \sqrt{(\# \text{matches})}$ if there are matches |
| OMIM gene | 1.0 if in OMIM gene | 1.0 if in OMIM gene |
| gnomAD allele frequency (overall) | 1.0 if not in gnomAD<br>0.5 if below 0.01% | 1.0 if not in gnomAD<br>0.5 if below 0.01% |
| HGMD pathogenicity | 0.5 if flagged as pathogenic<br>0.3 if flagged as likely pathogenic | 0.5 if flagged as pathogenic<br>0.3 if flagged as likely pathogenic |
| ClinVar pathogenicity | 1.0 if flagged as pathogenic<br>0.5 if flagged as likely pathogenic | 1.0 if flagged as pathogenic<br>0.5 if flagged as likely pathogenic |
| Conservedness (pyloP, 100 way) | 0.3 if bigger than 1.6 | 0.3 if bigger than 1.6 |
| Gene gnomAD o/e LoF | 0.5 if below 0.1 | 0.5 if below 0.1 |
| Gene inheritance mode | 0.5 if AD or XLD | 0.5 if AR or XLR |
| In-house database count (heterozygous) | 1.0 if below 3<br>0.5 if below 6 | - |
| Genotype | - | 1.0 if homozygous or two heterozygous hits in gene |

The top 10 candidates of each of the described models were then submitted to the LLM GPT-4o for inference of genotype-phenotype associations. Finally, the information from all models was transformed into features for the aiDIVA-meta model.

All models and competing approaches were benchmarked using 3,041 RD cases previously solved by genetics experts without using AI.

### Cohort Characteristics

The in-house cohort for benchmarking aiDIVA and competing classification models included 3,041 NGS datasets from affected individuals with RDs, including 1,975 ES and 1,066 GS datasets. This cohort comprised individuals presumed to be affected by monogenic disorders, each harboring either a single pathogenic variant (homozygous or heterozygous) or two compound heterozygous pathogenic variants in the corresponding disease-associated gene.

Cases with causal copy number variants (CNVs) or other types of structural variants (SVs) were excluded. NGS data processing, including alignment, variant calling, and variant annotation, was performed using the megSAP pipeline^1,37^. Causal variants were initially identified by genetics experts for diagnostic purposes at our institution using the clinical decision support system GSvar^37^.

Available clinical data included the disease group, observed disease phenotypes characterized by HPO terms, age at diagnosis, and gender. The cohort included 1,551 female, 1,444 male, and 46 undefined individuals. The age at diagnosis ranged from newborn to a maximum of 91 years (mean: 26.54 years, median: 21 years). The largest disease groups were mental, behavioral, or neurodevelopmental disorders (n=983); diseases of the visual system (n=597); diseases of the nervous system (n=587); neoplasms (n=301); endocrine, nutritional, or metabolic diseases (n=115); developmental anomalies (n=107); and diseases of the musculoskeletal system or connective tissue (n=90), as shown in **Supplemental Figure 2**.

### Pathogenicity Scoring

We trained the RF model aiDIVA-RF using 155,605 pathogenic and benign variants from ClinVar and 21 genomic features (**Supplemental Table 2**). Feature importance, quantified as the mean decrease in impurity, is shown in **Figure 2a**. Established pathogenicity estimators (e.g., AlphaMissense and REVEL), population allele frequency (gnomAD), evolutionary conservation (e.g., phyloP and phastCons), and predicted functional impact on proteins (i.e., the HIGH impact class of VEP) were among the most influential features contributing to accurate pathogenicity classification.

**Figure 2.**
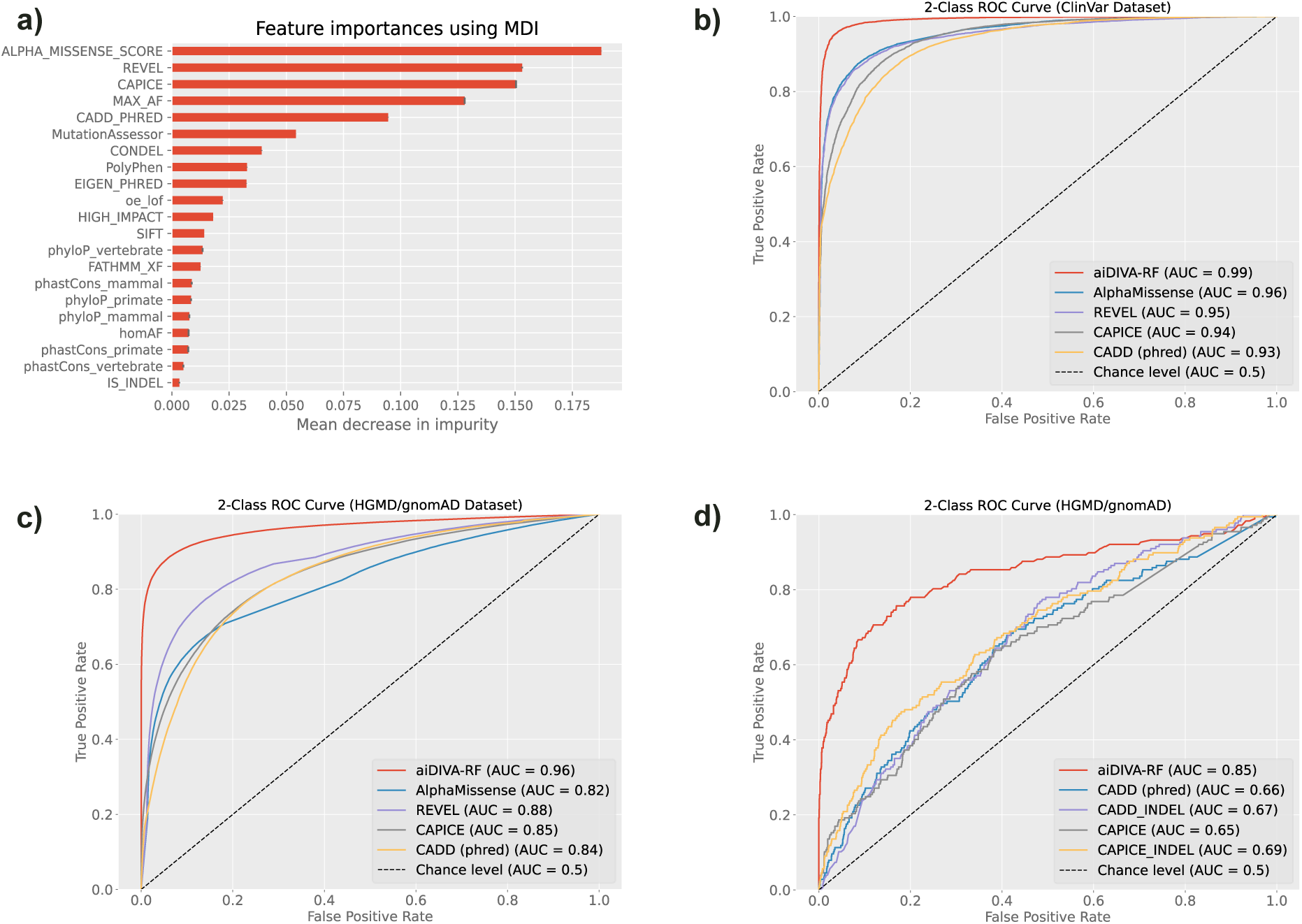
Pathogenicity Scoring **a)** Feature importances of the aiDIVA-RF pathogenicity model measured with the mean decrease in impurity (MDI) **b)** aiDIVA-RF pathogenicity prediction performance on ClinVar test set. **c)** aiDIVA-RF pathogenicity prediction performance on independent HGMD/gnomAD test set. **d)** Comparison between aiDIVA-RF pathogenicity model and two InDel specific pathogenicity scores.

We compared the performance of aiDIVA-RF for SNV classification with other pathogenicity or functional impact classifiers, including AlphaMissense^19^, REVEL^12^, CAPICE^11^, and CADD^17,18^. **Figure 2b** illustrates the performance of the models on the ClinVar test set with the respective receiver operating characteristic (ROC) area under the curve (AUC). To test the modeĺs robustness and generalizability, we conducted additional benchmarking using an independent validation dataset (**Figure 2c**) which included pathogenic variants sourced from HGMD and benign variants derived from gnomAD (see Methods). In both benchmarking analyses, aiDIVA-RF outperformed all competing methods. Specifically, when compared to AlphaMissense, aiDIVA-RF reached a ROC AUC of 0.99 versus 0.96 on the ClinVar test set and an ROC AUC of 0.96 versus 0.82 on the HGMD/gnomAD test set.

aiDIVA-RF incorporates a specialized model for in-frame indels, which simulates the impact of indels using a cluster of adjacent SNVs (see Methods). The performance of the indel pathogenicity classification could only be compared to CADD-indel and CAPICE-indel, as REVEL and AlphaMissense do not support indels. On the independent HGMD/gnomAD benchmark analyses, aiDIVA-RF demonstrated superior performance, achieving a ROC AUC of 0.85 (**Figure 2d**).

### Benchmarking Causal Variant Prioritization

aiDIVA provides two models for causal variant prioritization, which involve ranking the variants based on the likelihood of causing the genetic disease in the affected individual (aiDIVA-RF and aiDIVA-EB). A comparison of both approaches against Exomiser and AI-MARRVEL on a benchmark dataset of 3,041 expert-solved RD cases revealed the percentage of cases in which the models placed the correct variant on rank 1, up to rank 2, up to rank 3, and so on (**Figure 3**).

**Figure 3.**
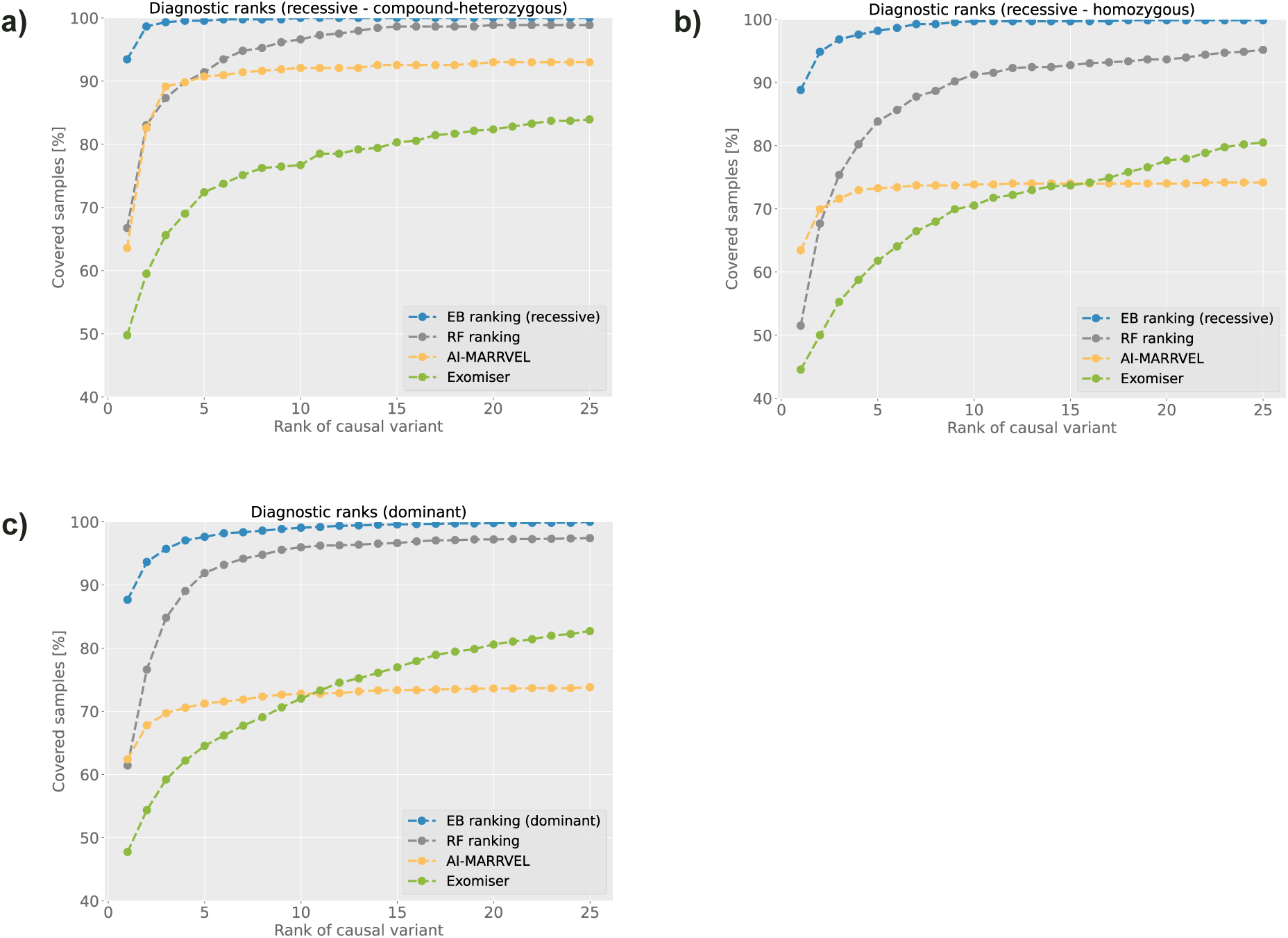
Diagnostic rank analysis comparison between the aiDIVA-RF ranking, the aiDIVA-EB-dominant/aiDIVA-EB-recessive ranking and two competing tools (Exomiser and AI-MARRVEL). We separated the results into three subsets based on the inheritance mode of the causal variants. **a)** Recessive - compound-heterozygous subset of our benchmark set (442 cases), **b)** recessive - homozygous subset (662 cases), **c)** dominant subset (1937 cases).

The benchmark set was split according to the mode of inheritance into the subsets compound heterozygous (**Figure 3a**), recessive (**Figure 3b**), and dominant (**Figure 3c**). aiDIVA-EB outperformed the other models in all inheritance modes, with approx. 90% of causal variants placed on rank 1. However, this level of performance is only anticipated in cases involving variants that have already been documented in at least one clinical database (hence referred to as the “evidence model”). aiDIVA-RF was superior across all inheritance modes compared to the other methods (AI-MARRVEL and Exomiser) not incorporating information from ClinVar, HGMD or similar clinical variant databases as features. The performance gains of aiDIVA were most notable for dominant inheritance (**Figure 3c**) and least evident for compound heterozygous traits (**Figure 3a**). AI-MARRVEL exhibits an unusual performance profile, achieving relatively high accuracy at the top-1 ranking level (50%–60%), but showing an early plateau at a comparatively low maximum (e.g., 73% for dominant disorders), suggesting limited sensitivity in broader candidate prioritization. Interestingly, Exomiser, a method published 10 years before AI-MARRVEL, still performed better than AI-MARRVEL for dominant and recessive cases when considering the top-20 ranked variants.

### Refining the Causal Variant Ranking Using Large Language Models

aiDIVA uses the LLM GPT-4o^33^ to select and explain the three most likely causal variant candidates out of the 10 highest ranked variants of each preceding model (aiDIVA-RF, aiDIVA-EB-dominant and aiDIVA-EB-recessive, **Figure 1**). The prompt for GPT-4o included affected genes, disease phenotypes (encoded as HPO terms), age at diagnosis, and gender (see Methods). We examined the LLM’s inference results using aiDIVA-RF rankings as input. Specifically, we focused on the 2,890 (out of 3,041) benchmark cases in which the causal variant was ranked within the top 10 by aiDIVA-RF. This subset was further stratified into two groups: an “easy” subset, comprising 2,528 cases where aiDIVA-RF ranked the causal variant within the top 3, and a “difficult” subset, consisting of the remaining 362 cases where the causal variant was ranked lower than third.

Initially, we tested a prompt version not specifying the individual variants affecting the top-10 genes. This approach yielded generally robust results, with rankings either improving or being stable in most cases. Notably, within the difficult subset, the ranking of the causal gene improved substantially, with 82.60% of cases (299 out of 362) now appearing in the top 3 reported by the LLM. However, in the easy subset, a decline in performance was observed in 7.63% of cases (193 out of 2,528), where the LLM no longer ranked the causal gene within the top 3. We hypothesized that 1) the reduced performance of the LLM in seemingly easy cases may be attributable to insufficient input information, and 2) cases with worsened rankings could be affected by limited reproducibility of LLM outputs.

We tested the reproducibility hypothesis by re-running the exact same prompt for two groups of cases from the “easy” group: 1) cases showing the correct gene among the top-3, and 2) cases with declined ranking not showing the correct gene in the top-3 as answer to the first prompt. While the results in group 1 remained almost the same in the second prompting of GPT-4o, with only one case worsening. The answers in group 2 (declined ranking in first answer) improved in 27.46 % of the cases (53/193), now reporting the correct variants among the top-3 as answers to the repeated identical prompt. We concluded that the reproducibility is lower in cases where the LLM struggles to find the correct answer. Hence, running each prompt twice (or several times) can be used to inform users about the robustness of the inference.

Furthermore, we tested the inclusion of additional information in the prompt, namely, the VEP impact (high, medium, or low) of each variant on the candidate genes, using the same two subgroups (improved/stable vs. declined ranking) of the “easy” cases. Inclusion of the variant effects in the prompt’s candidate gene list led to a strong performance gain in the declined-ranking group 2, with 40.93 % of causal variants (79/193) now among the top-3. The Inclusion of variant zygosity led to an additional mild improvement to 45.6 % causal variants (88/193) ranking among the top-3 (including the improvement from variant impact), while rankings in group 1 remained stable.

In summary, the enhanced ranking refinement by GPT-4o led to an improved variant prioritization, with 5 % more causal variants found among the top-3 ranked genes compared to using aiDIVA-RF only.

### Diagnostic Variant Ranking Using the aiDIVA-meta Model

To generate the final variant ranking suitable for clinical reporting, we developed an ensemble ML model named aiDIVA-meta, which integrates the outputs of all preceding models using the RF method (see Methods and **Figure 1**). To obtain realistic performance estimates for both know pathogenic variants (i.e. already reported in ClinVar or similar databases), as well as for novel variants or variants of unknown significance, we trained two versions of the ensemble: one incorporating all preceding models (aiDIVA-meta) and another excluding the evidence-based models (aiDIVA-EB-dominant and aiDIVA-EB-recessive), referred to as aiDIVA-meta-RF. As shown in **Figure 4a**, aiDIVA-meta and aiDIVA-meta-RF outperformed the individual ranking approaches. The aiDIVA-meta model (red curve) achieved the highest overall performance, ranking 97 % of the causal variants among the top-3, and 88% of the causal variants at rank 1. Of note, aiDIVA-meta ranked twice as many causal variants at rank 1 compared to Exomiser (green line). aiDIVA-meta-RF (purple line) significantly improved the results compared to aiDIVA-RF (grey line), despite excluding prior knowledge from clinical variant databases, underscoring the benefit of ensemble integration even without evidence-based features.

**Figure 4.**
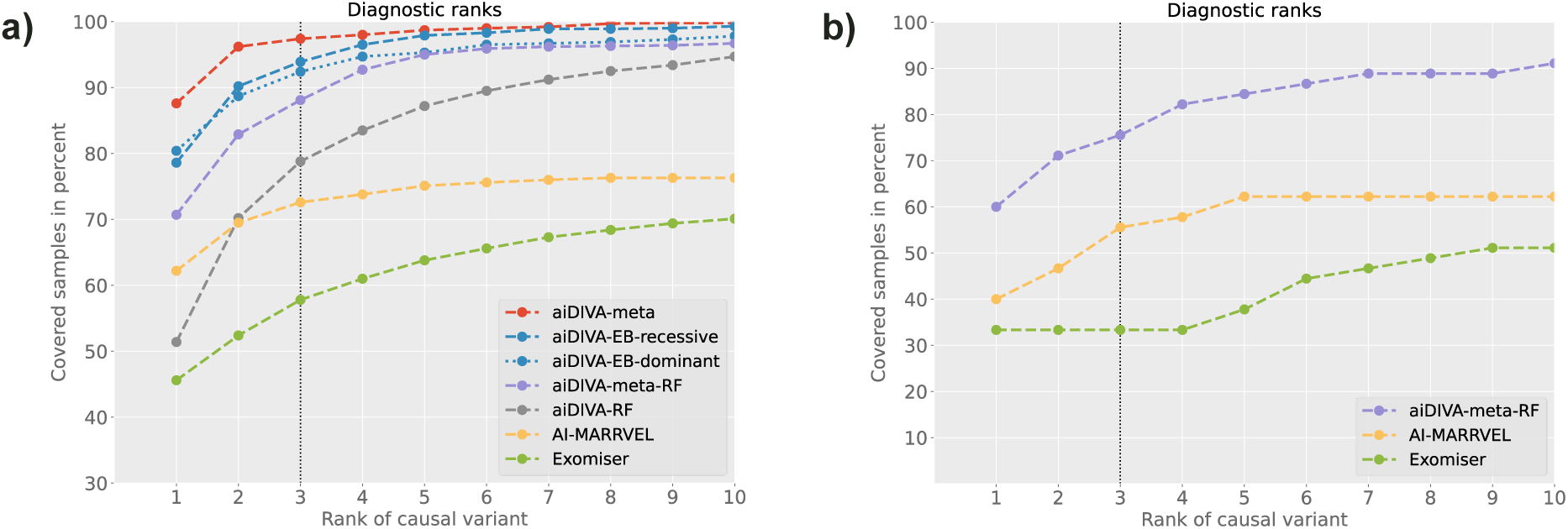
aiDIVA-meta performance. **a)** Diagnostic rank analysis comparison with the initial methods and the additional LLM refinement. The training for the aiDIVA-meta model was performed on 2/3 of our 3041 benchmark samples. This figure shows the ranking of the remaining 1/3 of the samples that were not used in the training. **b)** Diagnostic rank analysis comparison of 45 newly solved patients that were previously unsolvable. We excluded the evidence-based model here because it was initially used to prefilter the unsolved cases to find suitable cases for the manual reanalysis.

### Re-Analysis of Previously Unsolved Cases

To evaluate the utility of aiDIVA for the automated re-analysis of previously unsolved cases, we revisited 4,877 unsolved cases, originally evaluated using diagnostic-grade ES or GS prior to 2020. First, all cases were reannotated using the latest versions of relevant databases. Second, we prioritized 500 cases with the highest scoring top-ranked variants according to aiDIVA-EB, hypothesizing that newly available information (i.e., updated entries in ClinVar or HGMD) provide greatest opportunity of solving previously unsolved cases. A genetics expert then reviewed the selected 500 cases and identified 45 cases that were now considered to be solved (based on an ACMG classification of (likely) pathogenic). Finally, we assessed the ranking of these newly identified causal variants using Exomiser, AI-MARRVEL, and aiDIVA-meta-RF. As none of these tools used knowledge from ClinVar or HGMD, their performance did not achieve 100% sensitivity, as expected (**Figure 4b**). Nevertheless, aiDIVA-meta-RF strictly outperformed other tools placing 41 of the 45 causal variants within the top-10 ranks (91.1%), including 34 variants in the top-3 (75.6%). These results support the use of aiDIVA for automated prioritization of previously unsolved cases that may now receive a firm diagnosis based on the incorporation of new knowledge and updated gene scores.

## Discussion

### Improving RD Diagnostics with LLMs and Ensemble AI

LLMs represent a novel and promising addition to the diagnostic toolbox for genome medicine. Despite limited benchmarking studies, our results demonstrate that LLMs meaningfully improve the prioritization of causal variants and genes and offer clinically relevant explanations for top candidates. However, naïvely including all coding variants into a prompt often leads to performance degradation due to information overload and irrelevant signals^48^. To mitigate this, we combined the LLM approach with conventional ML classifiers and weighted-sum models to pre-select plausible variants for LLM-based inferences. This ensemble strategy, implemented in the aiDIVA-meta model, enabled causal variants to be ranked within the top-3 in 97% of expert-solved RD cases and hence can reduce the manual workload for clinical experts.

### Estimating In-Frame Indel Pathogenicity

Although relatively rare, in-frame indels represent approximately 5% of all the pathogenic variants annotated in ClinVar. Generally, in-frame indels remain underrepresented in variant databases and are poorly supported by most functional impact predictors as existing approaches for ML-based pathogenicity classification mostly ignore this variant type. In our study, we addressed this gap by representing in-frame indels as clusters of SNVs surrounding the affected region. This approximation significantly improved the classification performance, increasing the ROC AUC from 0.67 (CADD-Indel) to 0.85 (aiDIVA-RF). Nevertheless, the model’s performance on in-frame indels remained lower than that on SNVs, likely due to the limited training data and the inherent limitations of the SNV simulation strategy. Future improvements could include leveraging 3D protein structure modeling (e.g., with AlphaFold 3^49^) to estimate the functional effects of indels more accurately, although such integration was beyond the scope of this study.

### LLMs in Reanalysis of Unsolved Cases

Beyond initial diagnostics aiDIVA also proved valuable for re-analysis of previously unsolved cases. Reanalysis workflows typically involve a) periodic variant re-annotation b) scoring updates, and c) identification of cases with a strong increase in the top-ranked score for expert review^3,50–52^. LLMs can accelerate this process by generating explanations for newly prioritized variants, supporting expert triage. We demonstrated this by integrating aiDIVA into the genome-diagnostics platform megSAP, performing automated re-annotation, re-scoring, re-ranking of variants, and prioritization of solvable cases. In our retrospective analysis of 4,877 unsolved cases, aiDIVA preselected 500 high-confidence candidates based on updated scores, leading to 45 new diagnoses (approx. 1%) upon expert review. This demonstrates aiDIVA’s potential in large-scale automated reanalysis as variant databases evolve. Considering the rapid growth of genome datasets for RDs and still >60% unsolved cases, we expect a growing impact of the reanalysis model in clinical diagnostics.

### Limitations and Practical Considerations

While effective, LLM integration is not without limitations. Our benchmarking revealed that, in a small subset of cases, LLMs failed to reproduce answers consistently when the same prompt was repeated, indicating reduced reproducibility in borderline cases. Hence, the reproducibility of the answers can be used as a quality measure prior to making decisions. Moreover, new reasoning LLMs (for example GPT-o3^53^) can re-evaluate their own results, which is expected to increase the reproducibility of answers at the cost of longer computation times^54^.

We observed that including variant-level metadata, such as zygosity and functional impact, improved LLM performance in difficult cases. However, privacy concerns arise when sensitive patient data are included in prompts, especially when using cloud-based models. To address these concerns, one can either anonymize the information in the prompt (e.g., excluding variant positions) or set up local LLMs, which requires comprehensive expertise in AI.

Moreover, LLMs often fail to recognize that a diagnostic query is unsolvable with current knowledge. We observed that LLMs provide unreliable answers for cases that remained without definite diagnosis based on expert review. In some cases, GPT-4o returned references to nonexistent or irrelevant literature. These limitations highlight the need for additional safety measures, such as reproducibility checks or the integration of reasoning LLMs (e.g., GPT-o3), which can reevaluate and validate their own outputs at the cost of increased computation time.

Finally, structural variants, which account for a subset of genetic diagnoses, are not yet supported by *aiDIVA*, whereas Exomiser (since version 13.0.0) began to address this gap^55^.

### Model Specialization and LLM Enhancements

Current LLMs are trained on a very broad range of data but are not tailored for clinical diagnostics. However, they can be specialized using techniques such as fine-tuning on domain-specific data. Fine-tuning allows the specialization of pre-trained open-source LLMs to a knowledge domain by changing the weight of the upper neuron layers or by adding an additional layer^56^. It is performed by providing prompts with correct answers for a knowledge domain. For instance, cases with their phenotypic description and a list of genes/variants are provided as prompts, and the name of the causal gene is provided as the answer. Prior studies suggest that fine-tuning with as few as 1,000 domain-specific examples can yield substantial improvements in accuracy and contextual reasoning^57,58^. In addition, Retrieval Augmented Generation (RAGs) enables the embedding of external data sources, such as ClinVar or HPO, into the LLM^59^. Finally, refined strategies to pre-select the variants for LLM-inference could further improve the results, as would the inclusion of additional variant types, such as SVs. Ultimately, a switch to novel reasoning LLMs promises a strong gain in performance, reproducibility, and explainability of diagnostic AI.

### Future Directions

While aiDIVA demonstrates robust performance and clinical utility, several avenues remain for future development:

- Integration with Electronic Health Records and Clinical Feedback Loops: A major opportunity lies in integrating aiDIVA with electronic health record (EHR) systems. Direct access to structured phenotype data, longitudinal clinical notes, and family histories could support real-time re-analysis and continuous diagnostic refinement. Additionally, establishing a feedback loop in which clinicians validate or adjust aiDIVA’s suggestions would enable adaptive learning over time. This “human-in-the-loop” approach could improve model accuracy and contextual understanding, fostering transparency in clinical settings.
- Federated Learning and Privacy-Preserving Genomic AI: Given the sensitive nature of genomic data, privacy-preserving methods such as federated learning present an attractive avenue for enhancing aiDIVA without compromising patient confidentiality. Federated learning enables decentralized model training across institutions, where only model parameters, and not patient data, are shared. This approach would allow aiDIVA to benefit from diverse, multi-ethnic datasets while complying with data protection regulations, ultimately improving model robustness and generalizability.
- Multi-Omics Integration for Improved Variant Interpretation and Newborn Screening: Integrating multi-omics data into variant interpretation frameworks offers the potential to enhance diagnostic precision, particularly for variants of uncertain significance (VUS) or those located in non-coding or regulatory regions. Transcriptome data can help to identify aberrant splicing, allele-specific expression, or transcriptional dysregulation linked to specific variants. Likewise, proteomic and metabolomic data may provide downstream functional evidence, enabling better discrimination between benign and pathogenic alterations. This multi-layered evidence is especially relevant in newborn screening (NBS), where early detection of genetic disorders in still presymptomatic newborns is critical for timely intervention. As GS is increasingly being considered for use in NBS programs, AI-assisted interpretation tools, such as aiDIVA, will be essential to manage the large number of variants detected and provide clinically actionable insights.

In summary, aiDIVA provides AI models for assisting clinical experts with a highly sensitive and precise selection of potentially causal genetic variants in RDs. Besides ranking the correct causal variant among the top-3 positions in 97% of the 3,041 benchmarked cases, aiDIVA also returns explanations for these suggestions, which can be utilized for clinical report generation.

## Methods

### Model Overview

aiDIVA is a clinical decision support system for the classification, clinical interpretation and prioritization of likely clinically relevant variants observed in individuals affected with RDs. It comprises an ensemble of ML and statistical models, as shown in **Figure 1**. aiDIVA performs the following subsequential tasks to get a final variant ranking.

1. A RF model (aiDIVA-RF) is used to score and rank genetic variants according to their impact on protein function, potential pathogenicity, and fit to the respective phenotypes.
2. An evidence-based ranking model (aiDIVA-EB) is applied, which relies on known evidence for variant pathogenicity found in publications and disease variant databases (ClinVar, HGMD, and OMIM, etc.). aiDIVA-EB provides two separate models for recessive and dominant inheritance (aiDIVA-EB-dominant and aiDIVA-EB-recessive). Both models are based on a scoring function utilizing known evidence, as well as molecular-genetic features and phenotypic characteristics of the respective individual (see **Table 1** for a complete list).
3. The ten highest-ranking variants of the models aiDIVA-RF, aiDIVA-EB-dominant, and aiDIVA-EB-recessive are used as inputs for the LLM GPT-4o. GPT-4o is used to refine the rankings and to generate explanations for the three most likely causal variants, including references to scientific literature.
4. The top-10 variants of aiDIVA-RF, aiDIVA-EB-dominant, aiDIVA-EB-recessive, and the top-3 genes of their corresponding LLM refinement (LLM-aiDIVA-RF, LLM-aiDIVA-EB-dominant, and LLM-aiDIVA-EB-recessive) are combined as inputs for aiDIVA-meta, which creates the final pathogenicity ranking of an individual’s genetic variants.

An exemplary flowchart for variant annotation, pathogenicity ranking, and phenotype-based prioritization of the aiDIVA-RF model is provided in **Supplemental Figure 1**.

### Generating a Benchmarking Set for Diagnostic AIs

From 21,763 RD patients analyzed by GS or exome sequencing (ES) until 2023/06/01 at our institute, 4,186 cases received a firm genetic diagnosis. From the solved cohort, we extracted a benchmark set for diagnostic AIs based on the following criteria: 1) definite diagnosis, 2) monogenic cause, 3) one or two causal SNVs or indels with ACMG^5^ classification: pathogenic or likely pathogenic variant(s), and 4) causal coding SNVs or indels with a population allele frequency of less than 1%. Compound heterozygotes were included if at least one of the two causal variants was a coding SNV or indel. Cases with pathogenic structural, splicing or regulatory variants as well as repeat expansions were excluded from this benchmark dataset. This led to a final benchmark set of 3,041 clinically solved RD cases matching all criteria.

### Solving the Unsolved

To benchmark the ability of aiDIVA to increase the diagnostic yield via automated re-analysis, we used datasets from 4,877 cases analyzed in 2020 or earlier and without a firm diagnosis. The data were re-analyzed, and variants were re-annotated using the latest version of the diagnostic NGS-pipeline megSAP^36^. Based on the aiDIVA-EB ranking we then selected the top 500 cases based on the highest score for manual inspection by a clinical expert, resulting in 45 newly solved cases. These cases were used as a benchmark to test the ability of the AI models to solve unsolved cases.

### NGS Analysis and Variant Annotation with megSAP and Variant Effect Predictor

GS or ES data of clinical RD cases included in the benchmark sets described above were analyzed using the megSAP pipeline^36^, as described previously^1,37^. In brief, megSAP performs adapter trimming (SeqPurge), read alignment (BWA-mem2^38^), duplicate-flagging (SAMBLASTER^39^), indel-realignment (ABRA2^40^), and small variant calling (freebayes^41^). Part of the samples were analyzed using the Illumina DRAGEN^42^ (v4.3) mode of megSAP instead of the default pipeline described above. After variant calling, variants are annotated using a comprehensive in-house database combined with Variant Effect Predictor^43^. Variants were interpreted and classified according to the ACMG criteria^5^. Variant interpretation, variant filtering, and causal variant prioritization were performed by genetics experts and clinicians using the clinical decision support system GSvar^44^, as described previously^37^.

### ML-based Pathogenicity Estimation for Single Nucleotide Variants

We trained a RF model (aiDIVA-RF) to score the pathogenicity of genetic variants using a diverse set of genomic annotations as features. aiDIVA-RF has been implemented in Python using the package scikit-learn^45^. Variants for training and testing of aiDIVA-RF were obtained from the ClinVar database (2023/07/17). ClinVar comprises variants annotated during clinical testing, ranging from benign to pathogenic variants, and provides the respective disease phenotype in case of pathogenic or likely pathogenic variants^13^. To generate binary labeled data, we classified (likely) benign variants as negative and (likely) pathogenic variants as positive. ClinVar variants also present in the in-house benchmark sets (see above) were removed from the training data to prevent biases in the evaluation and comparison with other models. Furthermore, we removed non-coding, synonymous coding, splicing, and frameshift indel variants from the training/testing set, while retaining non-synonymous coding SNVs and in-frame indels. Moreover, variants in low-confidence regions, as defined in the megSAP pipeline at https://github.com/imgag/megSAP/blob/master/data/misc/low_conf_regions.bed, were removed to improve the quality of the training data. Finally, we removed variants with a minor allele frequency (MAF) of > 1% in gnomAD (v3.1.2).

The final ClinVar training dataset consisted of 90,400 (likely) pathogenic variants (SNVs: 87,750; in-frame indels: 2,650) and 65,202 (likely) benign variants (SNVs: 63,900; in-frame indels: 1,302). The dataset was split in a 90:10 ratio into independent training and testing datasets.

To investigate the generalizability of the trained RF model, we created a second test set using variants from the Human Gene Mutation Database (HGMD) (PRO_2022_04) not overlapping with ClinVar. Since HGMD only contains (likely) pathogenic variants, we included random likely benign variants from the gnomAD database (v3.1.2) with MAFs ranging from 0.1% to 1%, matching the upper limit in HGMD. We applied the same filters to the HGMD test data as described for the ClinVar data.

We used VEP to annotate all variants with various features (**Supplemental Table 2**), including maximum MAF, functional impact, evolutionary conservation, and loss-of-function intolerance. aiDIVA-RF was then trained using the ClinVar training set with binary labels (benign and pathogenic). The parameters used for training are listed in **Supplemental Table 5**.

### ML-based Pathogenicity Estimation for Small Insertions and Deletions

The annotation of in-frame indels remains difficult because they are not scored at all by many pathogenicity predictors or because pre-calculated scores are missing for them. To overcome this annotation issue, we converted both deletions and insertions into a cluster of subsequent SNVs, starting 3nt upstream and ending 3nt downstream of the indel. Positions modified in this manner were assigned a random nucleotide substitution. For deletions, all nucleotides along the deleted sequence and 3nt upstream/downstream were modified, whereas for insertions only the 3nt upstream and downstream were changed. Our procedure assumes that in-frame deletions and insertions affect their immediate neighborhood and surrounding regions but have a negligible effect on other regions of the protein. This strategy enabled the annotation of indels using the same feature set applied to SNVs. For each annotation feature, we computed the mean value across all random substitutions spanning the indel region. To incorporate in-frame indels in the aiDIVA-RF model, we added 2,650 pathogenic and 1,302 benign indels from ClinVar - using a 90:10 split for training and testing.

Frameshift indels are protein truncating and are therefore not included in aiDIVA-RF training. Instead, we annotated them as pathogenic with a score of 0.9 for subsequent steps.

### Prioritization of Causal Disease Variants Using Pathogenicity Scores and Phenotype Matching

Based on the predicted pathogenicity scores for non-synonymous SNVs and in-frame indels provided by aiDIVA-RF, we prioritized the most likely disease-causing variants using a ranking approach. Splice-variants were included in the ranking using precomputed SpliceAI scores. In addition to pathogenicity scores, aiDIVA uses HPO terms^25^ to score the similarity between phenotypes associated with a candidate gene and the respective disease phenotypes. Gene-associated HPO terms were derived from the HPO database (https://hpo.jax.org/, v2023-04-05). We used the Lin similarity measure to calculate the node-based semantic similarity between the two HPO term sets (gene-HPO set vs. patient-HPO set) within the hierarchical tree structure of the HPO ontology, as previously described^46^. The result of this calculation was used as a phenotype similarity score, indicating how well the patient’s phenotype matched a certain gene affected by a variant.

A second phenotype similarity score was computed by considering neighboring genes of the candidate disease gene based on protein-protein interaction information from STRING^47^ (v11.0b). If a gene interacting with the gene of interest had a high phenotypic similarity to the patient, calculated as described above, we assumed that the gene of interest may be an unknown cause of the disease.

A weighted sum approach was applied to combine the pathogenicity score, phenotype similarity, and gene network phenotype similarity using 2/3 of the solved in-house cases to train the weights. The training tested all weight combinations in 0.1 steps to minimize the rank of causal variants. To reduce the number of reported disease-variant candidates, variants with an aiDIVA-RF pathogenicity score below 0.7, which are also of low confidence, low complexity, or in repetitive regions, were removed. In addition, variants with a Lin score of zero for both phenotype similarity measures were excluded.

### Evidence-based Ranking

The evidence-based model aiDIVA-EB is a weighted sum-based ranking method developed for the clinical decision support system GSvar, which is routinely used for genome diagnostics. The input of aiDIVA-EB is an annotated list of variants and HPO-encoded individual phenotypes of a patient. Evidence-based ranking was conducted in two stages: (1) the exome/genome variant list was prefiltered to remove unlikely candidates; (2) the remaining variants were scored and ranked based on predefined weighted criteria.

1. We applied several filters: First, all variants with a MAF > 1% in gnomAD were excluded. Next, only variants in protein-coding and splicing regions were included, i.e. with a VEP impact of HIGH, MODERATE, or LOW. To remove pipeline-specific artifacts, we discarded variants that occurred more than ten times with the same genotype in an in-house database. Finally, variants with a base quality of less than 30 or a mapping quality of less than 20 were removed. After filtering, only rare coding and splicing variants unlikely to represent sequencing artifacts were retained. While some pathogenic variants have been reported in intronic and intergenic regions, these would typically be excluded by standard filtering criteria. To address this limitation, we retained variants that met the following criteria: i) were described as (likely) pathogenic in ClinVar (2024/08/05) or HGMD (PRO_2024_02) or that were predicted to affect splicing (SpliceAI > 0.5), and ii) had MAF < 1%. Typically, 200 – 500 variants remained after filtering.
2. These variants were scored using a point-based system, with separate models for dominant (aiDIVA-EB-dominant) and recessive (aiDIVA-EB-recessive) inheritance. Within each model, criteria were assessed independently, and points were assigned and summed up. Finally, the variants were ranked according to their overall scores in descending order. **Table 1** outlines the evidence criteria and corresponding point assignments for both aiDIVA-EB-dominant and aiDIVA-EB-recessive models.

### LLM Model

We used the LLM GPT-4o^33^ developed by OpenAI to refine the rankings generated by aiDIVA-RF and aiDIVA-EB, and to generate explanations for the suggested causal gene variants. We integrated GPT-4o into aiDIVA using the OpenAI RESTful API (https://platform.openai.com/docs/api-reference, accessed: 2025/05/23). For all analyses presented in this manuscript, we used the snapshot *gpt-4o-2024-05-13* of the GPT-4o model. Briefly, we first selected the top 10 ranked genes from each of the aiDIVA-RF, aiDIVA-EB-dominant, and aiDIVA-EB-recessive models as inputs for three separate queries (LLM-aiDIVA-RF, LLM-aiDIVA-EB-dominant, and LLM-aiDIVA-EB-recessive). Furthermore, each variant was assigned to one of five impact types (protein truncating, missense, in-frame insertion, in-frame deletion, and splice region). If the variant was present on both alleles, we added the keyword “homozygous”. In addition, we used all recorded disease phenotypes, age at diagnosis, and gender as characteristics to describe a case to GPT-4o. Combining this data, we automatically generated the following prompt for the respective individual in our in-house cohort (see also the Supplemental Section *Prompt Scheme*):

*“A [gender] rare disease patient of age [age] has the following symptoms: [list of phenotypes as HPO terms]. A causal variant in which of the following candidate genes would best explain these symptoms? For each candidate gene the consequence of the variant is given in brackets. Furthermore, homozygous variants are labeled in brackets. Candidate genes:*
*Gene-1 (loss-of-function variant)*
*Gene-2 (homozygous missense variant)*
*Gene-3 (splice region variant)*
*…*
*Gene-10 (homozygous inframe insertion/deletion)”*

To perform the analyses, we initialized the API using a comprehensive set of instructions (Supplemental Section *System Instructions*). Each query consisted of instructions and prompts (diagnostic questions) for five patients. Each of the five prompts were created according to the schema described above. As a response, we obtained a table according to the schema described in the system instructions given with each request. We requested reasoning from GPT-4o for choosing the 1^st^, 2^nd^, and 3^rd^ ranked genes, which can later be used to automatically generate diagnostic reports, if deemed appropriate by a genetics expert or a clinician.

### aiDIVA-meta

The aiDIVA-meta model is an ensemble of all ML and statistical methods presented above, which generates the final candidate disease variant/gene ranking. aiDIVA-meta again utilizes a RF model that was trained using the ranks and scores of all approaches (rankings generated by aiDIVA-RF, aiDIVA-EB-dominant, and aiDIVA-EB-recessive, LLM-aiDIVA-RF, LLM-aiDIVA-EB-dominant, and LLM-aiDIVA-EB-recessive) as features (**Supplemental Table 6**). To train aiDIVA-meta, we used 2/3 of our in-house benchmark set of 3,041 clinically solved Mendelian disease cases as training data and tested its performance on the remaining 1/3 of the cases. The training parameters are shown in **Supplemental Table 7**. The features and training parameters used for the aiDIVA-meta-RF model are shown in **Supplemental Table 8** and **Supplemental Table 9**.

### Benchmarking Exomiser

Exomiser is a variant prioritization tool used to rank candidate variants in patients with RDs^24^. It incorporates different pathogenicity scores and a random walk-based approach to measure the association between a gene’s and an individual’s disease phenotypes. For benchmarking, we used Exomiser v13.2.1. All parameters and settings applied are described in the Supplementary Section *Exomiser*. A detailed documentation of Exomiser usage is available at https://exomiser.readthedocs.io/en/13.2.1/ (accessed: 2025/05/23).

### Benchmarking AI-MARRVEL

AI-MARRVEL is a ML method for prioritizing causal variants in individuals with RDs^28^. Similar to aiDIVA and Exomiser, AI-MARRVEL integrates genomic and phenotypic information of affected individuals in their model. We used AI-MARRVEL v1.0.1 (https://github.com/LiuzLab/AI_MARRVEL, accessed: 2024/07/31) and ran the AI-MARRVEL Docker image with default settings to generate all benchmark results.

## Ethics Declaration

Written informed consent was obtained from all individuals or their guardians and archived. All procedures were performed in accordance with the Helsinki Declaration. Individual-level data were de-identified. The study was approved by the ethics committee of the medical faculty by the local Institutional Review Board of the Medical Faculty of the University of Tübingen, Germany (066/2021BO2).

## Conflicts of Interests

Daniela Bezdan, Stephan Ossowski, Marc Sturm and Tobias Haack are cofounders and shareholders of dxOmics GmbH.

The other authors declare no competing interests.

## Data Availability

The aiDIVA-EB models are available via GSvar^44^ and through the command line tool VariantRanking of ngs-bits from https://github.com/imgag/ngs-bits/blob/master/doc/tools/VariantRanking/index.md with model names GSvar_v2_dominant and GSvar_v2_recessive.

The source code of aiDIVA, as well as all three trained random forest models (aiDIVA-RF, aiDIVA-meta and aiDIVA-meta-RF) are available in the GitHub repository at https://github.com/imgag/aiDIVA and through the following Zenodo record https://zenodo.org/records/16966353.

## Acknowledgement

First and foremost, all authors thank the families for participating in the study. T.B.H received funding from the European Commission (Recon4IMD - GAP-101080997) and the German Research Foundation (DFG; research grant numbers 418081722, 433158657, EJP-RD Artemis: 542553983). S.O. and D.Be. were supported by the European Union via the project European Rare Disease Research Alliance (ERDERA), GA n°101156595, funded under call HORIZON-HLTH-2023-DISEASE-07 and the European Union’s Horizon 2020 research and innovation programme under grant agreement No 779257 (Solve-RD). S.O. received funding from the German Research Foundation (DFG; research grant numbers 514060894, 514208594, 514177729, and OS 647/1-1).

## References

1. Riess O, Sturm M, Menden B, et al. Genomes in clinical care. Npj Genomic Med. 2024;9(1):20. doi:10.1038/s41525-024-00402-2

2. Best S, Fehlberg Z, Richards C, et al. Reanalysis of genomic data in rare disease: current practice and attitudes among Australian clinical and laboratory genetics services. Eur J Hum Genet. 2024;32(11):1428–1435. doi:10.1038/s41431-024-01633-8

3. Laurie S, Steyaert W, de Boer E, et al. Genomic reanalysis of a pan-European rare-disease resource yields new diagnoses. Nat Med. 2025;31(2):478–489. doi:10.1038/s41591-024-03420-w

4. Olfson E, Cottrell CE, Davidson NO, et al. Identification of Medically Actionable Secondary Findings in the 1000 Genomes. PLOS ONE. 2015;10(9):e0135193. doi:10.1371/journal.pone.0135193

5. Richards S, Aziz N, Bale S, et al. Standards and guidelines for the interpretation of sequence variants: a joint consensus recommendation of the American College of Medical Genetics and Genomics and the Association for Molecular Pathology. Genet Med. 2015;17(5):405–423. doi:10.1038/gim.2015.30

6. Melidis DP, Landgraf C, Schmidt G, et al. GenOtoScope: Towards automating ACMG classification of variants associated with congenital hearing loss. PLOS Comput Biol. 2022;18(9):e1009785. doi:10.1371/journal.pcbi.1009785

7. Munté E, Feliubadaló L, Pineda M, et al. vaRHC: an R package for semi-automation of variant classification in hereditary cancer genes according to ACMG/AMP and gene-specific ClinGen guidelines. Bioinformatics. 2023;39(3):btad128. doi:10.1093/bioinformatics/btad128

8. Adzhubei IA, Schmidt S, Peshkin L, et al. A method and server for predicting damaging missense mutations. Nat Methods. 2010;7(4):248–249. doi:10.1038/nmeth0410-248

9. Adzhubei I, Jordan DM, Sunyaev SR. Predicting Functional Effect of Human Missense Mutations Using PolyPhen-2. Curr Protoc Hum Genet. 2013;76(1):7.20.1–7.20.41. doi:10.1002/0471142905.hg0720s76

10. Rogers MF, Shihab HA, Mort M, Cooper DN, Gaunt TR, Campbell C. FATHMM-XF: accurate prediction of pathogenic point mutations via extended features. Bioinformatics. 2018;34(3):511–513. doi:10.1093/bioinformatics/btx536

11. Li S, van der Velde KJ, de Ridder D, et al. CAPICE: a computational method for Consequence-Agnostic Pathogenicity Interpretation of Clinical Exome variations. Genome Med. 2020;12(1):1–11. doi:10.1186/s13073-020-00775-w

12. Ioannidis NM, Rothstein JH, Pejaver V, et al. REVEL: An Ensemble Method for Predicting the Pathogenicity of Rare Missense Variants. Am J Hum Genet. 2016;99(4):877–885. doi:10.1016/j.ajhg.2016.08.016

13. Landrum MJ, Lee JM, Benson M, et al. ClinVar: improving access to variant interpretations and supporting evidence. Nucleic Acids Res. 2018;46(D1):D1062–D1067. doi:10.1093/nar/gkx1153

14. Stenson PD, Mort M, Ball EV, Shaw K, Phillips AD, Cooper DN. The Human Gene Mutation Database: building a comprehensive mutation repository for clinical and molecular genetics, diagnostic testing and personalized genomic medicine. Hum Genet. 2014;133(1):1–9. doi:10.1007/s00439-013-1358-4

15. Livesey BJ, Marsh JA. Variant effect predictor correlation with functional assays is reflective of clinical classification performance. Genome Biol. 2025;26(1):1–27. doi:10.1186/s13059-025-03575-w

16. Ionita-Laza I, McCallum K, Xu B, Buxbaum JD. A spectral approach integrating functional genomic annotations for coding and noncoding variants. Nat Genet. 2016;48(2):214–220. doi:10.1038/ng.3477

17. Rentzsch P, Witten D, Cooper GM, Shendure J, Kircher M. CADD: predicting the deleteriousness of variants throughout the human genome. Nucleic Acids Res. 2019;47(D1):D886–D894. doi:10.1093/nar/gky1016

18. Rentzsch P, Schubach M, Shendure J, Kircher M. CADD-Splice—improving genome-wide variant effect prediction using deep learning-derived splice scores. Genome Med. 2021;13(1):1–12. doi:10.1186/s13073-021-00835-9

19. Cheng J, Novati G, Pan J, et al. Accurate proteome-wide missense variant effect prediction with AlphaMissense. Science. 2023;381(6664):eadg7492. doi:10.1126/science.adg7492

20. Jumper J, Evans R, Pritzel A, et al. Highly accurate protein structure prediction with AlphaFold. Nature. 2021;596(7873):583-+. doi:10.1038/s41586-021-03819-2

21. Ferlaino M, Rogers MF, Shihab HA, et al. An integrative approach to predicting the functional effects of small indels in non-coding regions of the human genome. BMC Bioinformatics. 2017;18(1):1–8. doi:10.1186/s12859-017-1862-y

22. Jaganathan K, Kyriazopoulou Panagiotopoulou S, McRae JF, et al. Predicting Splicing from Primary Sequence with Deep Learning. Cell. 2019;176(3):535–548.e24. doi:10.1016/j.cell.2018.12.015

23. Jaganathan K, Ersaro N, Novakovsky G, et al. Predicting expression-altering promoter mutations with deep learning. Science. 2025;389(6760):eads7373. doi:10.1126/science.ads7373

24. Smedley D, Jacobsen JOB, Jäger M, et al. Next-generation diagnostics and disease-gene discovery with the Exomiser. Nat Protoc. 2015;10(12):2004–2015. doi:10.1038/nprot.2015.124

25. Köhler S, Gargano M, Matentzoglu N, et al. The Human Phenotype Ontology in 2021. Nucleic Acids Res. 2021;49(D1):D1207–D1217. doi:10.1093/nar/gkaa1043

26. Vestito L, Jacobsen JOB, Walker S, et al. Efficient reinterpretation of rare disease cases using Exomiser. Npj Genomic Med. 2024;9(1):65. doi:10.1038/s41525-024-00456-2

27. Bosio M, Drechsel O, Rahman R, et al. eDiVA—Classification and prioritization of pathogenic variants for clinical diagnostics. Hum Mutat. 2019;40(7):865–878. doi:10.1002/humu.23772

28. Mao D, Liu C, Wang L, et al. AI-MARRVEL — A Knowledge-Driven AI System for Diagnosing Mendelian Disorders. NEJM AI. 2024;1(5):AIoa2300009. doi:10.1056/AIoa2300009

29. Boudellioua I, Kulmanov M, Schofield PN, Gkoutos GV, Hoehndorf R. DeepPVP: phenotype-based prioritization of causative variants using deep learning. BMC Bioinformatics. 2019;20(1):1–8. doi:10.1186/s12859-019-2633-8

30. Bojić L, Kovačević P, Čabarkapa M. Does GPT-4 surpass human performance in linguistic pragmatics? Humanit Soc Sci Commun. 2025;12(1):794. doi:10.1057/s41599-025-04912-x

31. Kim J, Wang K, Weng C, Liu C. Assessing the utility of large language models for phenotype-driven gene prioritization in the diagnosis of rare genetic disease. Am J Hum Genet. 2024;111(10):2190–2202. doi:10.1016/j.ajhg.2024.08.010

32. McDuff D, Schaekermann M, Tu T, et al. Towards accurate differential diagnosis with large language models. Nature. 2025;642(8067):451–457. doi:10.1038/s41586-025-08869-4

33. OpenAI, Hurst A, Lerer A, et al. GPT-4o System Card. arXiv. Preprint posted online October 25, 2024. doi:10.48550/arXiv.2410.21276

34. Touvron H, Lavril T, Izacard G, et al. LLaMA: Open and Efficient Foundation Language Models. arXiv. Preprint posted online February 27, 2023. doi:10.48550/arXiv.2302.13971

35. Jiang AQ, Sablayrolles A, Mensch A, et al. Mistral 7B. arXiv. Preprint posted online October 10, 2023. doi:10.48550/arXiv.2310.06825

36. Sturm M, Admard J, Schütz L, et al. imgag/megSAP: 2025_03. Published online March 21, 2025. doi:10.5281/zenodo.15063428

37. Park J, Sturm M, Seibel-Kelemen O, Ossowski S, Haack TB. Lessons Learned from Translating Genome Sequencing to Clinical Routine: Understanding the Accuracy of a Diagnostic Pipeline. Genes. 2024;15(1):136. doi:10.3390/genes15010136

38. Vasimuddin Md, Misra S, Li H, Aluru S. Efficient Architecture-Aware Acceleration of BWA-MEM for Multicore Systems. In: 2019 IEEE International Parallel and Distributed Processing Symposium (IPDPS). 2019:314-324. doi:10.1109/IPDPS.2019.00041

39. Faust GG, Hall IM. SAMBLASTER: fast duplicate marking and structural variant read extraction. Bioinformatics. 2014;30(17):2503–2505. doi:10.1093/bioinformatics/btu314

40. Mose LE, Perou CM, Parker JS. Improved indel detection in DNA and RNA via realignment with ABRA2. Bioinformatics. 2019;35(17):2966–2973. doi:10.1093/bioinformatics/btz033

41. Garrison E, Marth G. Haplotype-based variant detection from short-read sequencing. arXiv. Preprint posted online July 20, 2012. doi:10.48550/arXiv.1207.3907

42. Behera S, Catreux S, Rossi M, et al. Comprehensive genome analysis and variant detection at scale using DRAGEN. Nat Biotechnol. 2025;43(7):1177–1191. doi:10.1038/s41587-024-02382-1

43. McLaren W, Gil L, Hunt SE, et al. The Ensembl Variant Effect Predictor. Genome Biol. 2016;17(1):1–14. doi:10.1186/s13059-016-0974-4

44. Sturm M, Chernov A, Schütz L, et al. imgag/ngs-bits: 2025_03. Published online March 19, 2025. doi:10.5281/zenodo.15051584

45. Pedregosa F, Varoquaux G, Gramfort A, et al. Scikit-learn: Machine Learning in Python. J Mach Learn Res. 2011;12(85):2825–2830.

46. Lin D. An Information-Theoretic Definition of Similarity. In: Proceedings of the Fifteenth International Conference on Machine Learning. ICML ’98. Morgan Kaufmann Publishers Inc.; 1998:296–304.

47. Szklarczyk D, Gable AL, Lyon D, et al. STRING v11: protein–protein association networks with increased coverage, supporting functional discovery in genome-wide experimental datasets. Nucleic Acids Res. 2019;47(D1):D607–D613. doi:10.1093/nar/gky1131

48. Li Z, Xie B, Hilsabeck R, et al. Effects of Different Prompts on the Quality of GPT-4 Responses to Dementia Care Questions. In: IEEE Computer Society; 2024:412–417. doi:10.1109/ICHI61247.2024.00059

49. Abramson J, Adler J, Dunger J, et al. Accurate structure prediction of biomolecular interactions with AlphaFold 3. Nature. 2024;630(8016):493–500. doi:10.1038/s41586-024-07487-w

50. Baker SW, Murrell JR, Nesbitt AI, et al. Automated Clinical Exome Reanalysis Reveals Novel Diagnoses. J Mol Diagn. 2019;21(1):38–48. doi:10.1016/j.jmoldx.2018.07.008

51. Deignan JL, Chung WK, Kearney HM, Monaghan KG, Rehder CW, Chao EC. Points to consider in the reevaluation and reanalysis of genomic test results: a statement of the American College of Medical Genetics and Genomics (ACMG). Genet Med. 2019;21(6):1267–1270. doi:10.1038/s41436-019-0478-1

52. Matalonga L, Hernández-Ferrer C, Piscia D, et al. Solving patients with rare diseases through programmatic reanalysis of genome-phenome data. Eur J Hum Genet. 2021;29(9):1337–1347. doi:10.1038/s41431-021-00852-7

53. OpenAI. OpenAI o3 and o4-mini. April 16, 2025. Accessed June 16, 2025. https://openai.com/index/introducing-o3-and-o4-mini/

54. Zhang M, Bjerva J, Biswas R. Scaling Reasoning can Improve Factuality in Large Language Models. arXiv. Preprint posted online May 16, 2025. doi:10.48550/arXiv.2505.11140

55. Jacobsen JOB, Kelly C, Cipriani V, et al. Phenotype-driven approaches to enhance variant prioritization and diagnosis of rare disease. Hum Mutat. 2022;43(8):1071–1081. doi:10.1002/humu.24380

56. Houlsby N, Giurgiu A, Jastrzebski S, et al. Parameter-Efficient Transfer Learning for NLP. In: Proceedings of the 36th International Conference on Machine Learning. PMLR; 2019:2790–2799. Accessed August 29, 2025. https://proceedings.mlr.press/v97/houlsby19a.html

57. Zhou C, Liu P, Xu P, et al. LIMA: Less Is More for Alignment. In: Proceedings of the 37th International Conference on Neural Information Processing Systems. NIPS ’23. Curran Associates Inc.; 2023:55006–55021.

58. Muennighoff N, Yang Z, Shi W, et al. s1: Simple test-time scaling. arXiv. Preprint posted online March 1, 2025. doi:10.48550/arXiv.2501.19393

59. Lewis P, Perez E, Piktus A, et al. Retrieval-Augmented Generation for Knowledge-Intensive NLP Tasks. arXiv. Preprint posted online April 12, 2021. doi:10.48550/arXiv.2005.11401

