## Supplemental Material for "aiDIVA – Diagnostics of Rare Genetic Diseases Using Large Language Models"

#### Annotation of genetic variants for random forest-based pathogenicity classification, ranking and causal variant prioritization

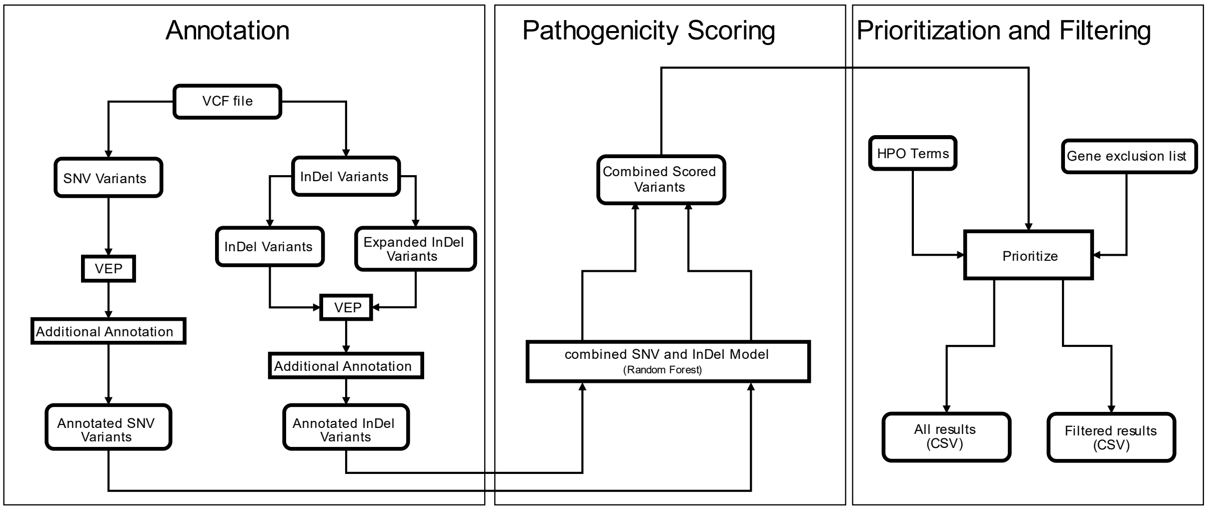


**Supplemental Figure 1** Flowchart for the usage of the trained RF model for pathogenicity classification and causal variant prioritization. A) Indels are transformed into SNV clusters, SNVs are annotated using Variant Effect Predictor, B) each variant is classified and assigned a score (0.0 - 1.0) by the RF model, C) the genomic ranking from the RF is combined with two phenotype-similarity measures (patient's phenotype vs. gene-associated phenotypes) to calculate a combined ranking. Results are exported to csv files allowing for additional filtering steps.

The workflow of the random forest-based ranking included in aiDIVA consists of three parts (see **Supplemental Figure 1**) with an initial filter step. The initial filtering is used to restrict the patient's variants to the coding part of the genome. This step is performed to reduce the computational effort to run aiDIVA-RF. Currently aiDIVA-RF only supports coding variants (see **Supplemental Table 1** for an overview of supported variant consequence terms). The *Annotation* part can be skipped if the data was already annotated beforehand. The annotation in aiDIVA-RF is performed with the variant effect predictor (VEP)[1] (version 109.3) for the basic annotation and ngs-bits[2] (https://github.com/imgag/ngs-bits) (version 2025_03) for additional annotation sources. The annotation of the variants covers the following features: The population allele frequencies for each variant based on the gnomAD genome dataset[3]. Evolutionary conservation information based on phyloP[4] and phastCons[5]. Damage scores that give information on the functional impact of a variant based on the following algorithms SIFT[6], PolyPhen[7], Condel[8], CADD[9], CAPICE[10], REVEL[11], MutationAssessor[12], FATHMM-XF[13] and Alpha Missense[14]. Genomic region-specific information on the variants including segmental duplications[15,16] and simple repeats[17] taken from the UCSC table browser[18] and RepeatMasker[19]. From the gnomAD gene constraints[20] the observed/expected loss of function score (oe_lof) is incorporated to give insights into the tolerance of loss of function variants in a gene. For splicing variants, we use the precomputed masked SpliceAI[21] scores (date of creation 2023/09/29). Furthermore, we calculated the homozygous allele frequency (homAF) based on the gnomAD genome database. The exact version of the beforementioned databases and tools can be found in **Supplemental Table 2**.

**Supplemental Table 1** Variant types included in the random forest-based ranking of aiDIVA.

| |
|---|
| start_retained_variant |

| |
|---|
| start_lost |
| synonymous_variant |
| missense_variant |
| inframe_insertion |
| inframe_deletion |
| stop_gained |
| frameshift_variant |
| coding_sequence_variant |
| splice_acceptor_variant |
| splice_donor_variant |
| splice_region_variant |
| stop_lost |
| stop_retained_variant |
| incomplete_terminal_codon_variant |
| protein_altering_variant |

**Supplemental Table 2** Genomic and population genetics characteristics used as features for the random forest model for variant pathogenicity classification.

| Feature | Type | Version / Date of creation |
|---|---|---|
| SIFT | Functional Impact | v6.2.1 |
| PolyPhen | Functional Impact | v2.2.3 |
| CADD (phred scaled score) | Functional Impact | v1.6 |
| REVEL | Functional Impact | V1.3 |
| gnomAD MAX_AF (Maximum Minor Allele Frequency) | Population Allele Frequency | gnomAD genome v3.1.2 |
| FATHMM_XF | Functional Impact | 2017/08/09 |
| MutationAssessor | Functional Impact | v3 |
| phastCons_mammal (30way) | Evolutionary Conservation | 2017/11/06 |
| phastCons_primate (17way) | Evolutionary Conservation | 2015/02/27 |
| phastCons_vertebrate (100way) | Evolutionary Conservation | 2015/05/08 |
| phyloP_mammal (30way) | Evolutionary Conservation | 2017/11/06 |
| phyloP_primate (17way) | Evolutionary Conservation | 2015/02/27 |
| phyloP_vertebrate (100way) | Evolutionary Conservation | 2015/05/08 |
| oe_lof | Tolerance to Inactivation | gnomAD v2.1.1 |
| homAF | Population Allele Frequency | gnomAD genome v3.1.2 |
| CAPICE | Functional Impact | v1 |
| Condel | Functional Impact | FannsDB v2.0 |
| Eigen (phred scaled score) | Functional Impact | v1.0.0 |
| AlphaMissense | Functional Impact | v1 |
| High impact (binary) | Functional Impact | Ensembl VEP 109.3 |
| is indel (binary) | Variant characteristic | not applicable |

**Supplemental Table 3** Filters applied during the prioritization of the random forest-based ranking.

| Filter | Description |
|---|---|
| Low_conf_regions | discard all variants that lie in a low confidence region (as defined in the megSAP pipeline[22]) |
| Repeatmasker | discard all variants that have a pathogenicity score of less than 0.7 and overlap with a region contained in RepeatMasker |

| | |
|---|---|
| Off-target | discard all variants that lie outside of the target region |
| Simple tandem repeats | discard all variants that have a pathogenicity score of less than 0.7 and overlap with a simple tandem repeat found in the UCSC genome browser |
| Segment duplication | discard all variants that have a pathogenicity score of less than 0.7 and overlap with a segment duplication found in the UCSC genome browser |
| Homopolymer region | discard indel variants with a pathogenicity score of less than 0.7 that lie in a homopolymer region |
| Low complexity region | discard indel variants with a pathogenicity score of less than 0.7 that lie in a low complexity region |
| Maximum Minor Allele Frequency threshold | discard variants that are more frequent than 1% in the population |
| HPO relatedness filter | discard variants that have no HPO relation at all to the given HPO set |
| Coding filter | discard all variants that are synonymous or non-coding (frameshifts and splicing variants are kept and handled separately) |

**Supplemental Table 4** All features used to train the random forest model sorted according to their importance.

| **Feature** | **Importance** |
|---|---|
| ALPHA_MISSENSE_SCORE | 0.187624 |
| REVEL | 0.153069 |
| CAPICE | 0.150424 |
| MAX_AF | 0.127964 |
| CADD_PHRED | 0.094500 |
| MutationAssessor | 0.054205 |
| CONDEL | 0.039225 |
| PolyPhen | 0.032817 |
| EIGEN_PHRED | 0.032590 |
| oe_lof | 0.022225 |
| HIGH_IMPACT | 0.018015 |
| SIFT | 0.014102 |
| phyloP_vertebrate | 0.013387 |
| FATHMM_XF | 0.012512 |
| phastCons_mammal | 0.008709 |
| phyloP_primate | 0.008346 |
| phyloP_mammal | 0.007581 |
| homAF | 0.007301 |
| phastCons_primate | 0.007122 |
| phastCons_vertebrate | 0.005051 |
| IS_INDEL | 0.003231 |

**Supplemental Table 5** Parameter settings used to train the random forest model aiDIVA-RF.

| **Parameter** | **Value** |
|---|---|
| bootstrap | True |
| ccp_alpha | 0.0 |
| class_weight | None |

| | |
|---|---|
| criterion | gini |
| max_depth | None |
| max_features | sqrt |
| max_leaf_nodes | None |
| max_samples | None |
| min_impurity_decrease | 0.0 |
| min_samples_leaf | 1 |
| min_samples_split | 2 |
| min_weight_fraction_leaf | 0.0 |
| n_estimators | 1000 |
| n_jobs | None |
| oob_score | False |
| random_state | 14038 |
| verbose | 0 |
| warm_start | False |

#### Exomiser

Parameters used for running Exomiser.
PathogenicitySources:

- REVEL
- MVP
- CADD

FrequencySources:

- GNOMAD_G_AFR
- GNOMAD_G_AMR
- GNOMAD_G_EAS
- GNOMAD_G_NFE
- GNOMAD_G_SAS

VariantEffectFilter:

- FIVE_PRIME_UTR_EXON_VARIANT
- FIVE_PRIME_UTR_INTRON_VARIANT
- THREE_PRIME_UTR_EXON_VARIANT
- THREE_PRIME_UTR_INTRON_VARIANT
- NON_CODING_TRANSCRIPT_EXON_VARIANT
- UPSTREAM_GENE_VARIANT
- INTERGENIC_VARIANT
- REGULATORY_REGION_VARIANT
- CODING_TRANSCRIPT_INTRON_VARIANT
- NON_CODING_TRANSCRIPT_INTRON_VARIANT
- DOWNSTREAM_GENE_VARIANT

Example Exomiser config file:

*## Exomiser Analysis – NA12878*

---

*analysis:*

*genomeAssembly: hg38*

*vcf: NA12878.vcf*

*ped:*

*proband: NA12878*

*hpoIds: [HP:xxxxxxx, HP:xxxxxxx]*

*analysisMode: PASS_ONLY*

*frequencySources: [GNOMAD_G_AFR, GNOMAD_G_AMR, GNOMAD_G_EAS, GNOMAD_G_NFE, GNOMAD_G_SAS]*

*pathogenicitySources: [REVEL, MVP, CADD]*

*steps: [*

*variantEffectFilter: {remove: [FIVE_PRIME_UTR_EXON_VARIANT, FIVE_PRIME_UTR_INTRON_VARIANT, THREE_PRIME_UTR_EXON_VARIANT, THREE_PRIME_UTR_INTRON_VARIANT, NON_CODING_TRANSCRIPT_EXON_VARIANT, UPSTREAM_GENE_VARIANT, INTERGENIC_VARIANT, REGULATORY_REGION_VARIANT, CODING_TRANSCRIPT_INTRON_VARIANT, NON_CODING_TRANSCRIPT_INTRON_VARIANT, DOWNSTREAM_GENE_VARIANT]},*

*frequencyFilter: {maxFrequency: 1.0},*

*pathogenicityFilter: {keepNonPathogenic: true},*

*omimPrioritiser: {},*

*phenixPrioritiser: {}*

*]*

*outputOptions:*

*outputContributingVariantsOnly: false*

*numGenes: 0*

*outputDirectory: results_exomiser/*

*outputFileName: NA12878_exomiser*

*outputFormats: [TSV-VARIANT]*

*...*

#### OpenAI RESTful API

##### System Instructions

To compile a query for the OpenAI API we combined a comprehensive set of general instructions with patient-specific prompts. Usually, a single query consists of the general instructions and prompts for 5 patients. Queries can contain any number of patient-specific prompts, however, queries with more than 20 patient-specific prompts are sometimes prematurely aborted by GPT-4o[23]. We used the following general instructions GPT-4o, which explain to GPT-4o the diagnostic setting of the query as well as the desired scope and output format of the answer:

*“Create a clinical expert in rare diseases that answers given prompts in the following way:*
*Please create table with 8 columns with one row for each question. Do answer every question independently, treat every question as a new, standalone question and answer without referring to previous interactions.*
*Write my complete prompt into the first column.*
*Now I want to explain you the table properties. The table has 8 columns.*
*Column 1 should have the name 'prompt' and should contain my entire question. Do not modify it.*
*Column 2 should have the name '1st ranked Gene' and should contain your 1st choice of the causal gene for the given phenotype.*
*Column 3 should have the name '1st ranked Gene explanation' and should contain the explanation for your 1st choice for causal gene. Please add the source for your answer in your text inside the table.*
*Column 4 should have the name '2nd ranked Gene' and should contain your 2nd choice of the causal gene.*
*Column 5 should have the name '2nd ranked Gene explanation' and should contain the explanation for your 2nd choice for causal gene. Please add the source for your answer in your text inside the table.*
*Column 6 should have the name '3rd ranked Gene' and should contain your 3rd choice of the causal gene.*
*Column 7 should have the name '3rd ranked Gene explanation' and should contain the explanation for your 3rd choice for causal gene. Please add the source for your answer in your text inside the table.*
*Column 8 should have the name 'source' and should contain all sources and bibliography of your sources you needed to answer this question. Please add sources in form of full web addresses within your answer in your text inside the table.*
*Please provide the result table in plain json.”*

#### Patient-specific Prompts

Following the general instructions, we include the following description of each interrogated patient in the prompt:

*“A [sex] rare disease patient of age [age] has the following symptoms: [list of phenotypes as HPO terms]. A causal variant in which of the following candidate genes would best explain these symptoms? For each candidate gene the consequence of the variant is given in brackets. Furthermore, homozygous variants are labeled in brackets. Candidate genes:*
*Gene-1 (loss-of-function variant)*
*Gene-2 (homozygous missense variant)*
*Gene-3 (splice region variant)*
*…*
*Gene-10 (homozygous inframe insertion/deletion):”*

#### Response in JSON format

The GPT-4o API returns the answer in the following JSON format:

```
[
  {
    "prompt": "<Request sent to the LLM>",
    "1st ranked Gene": "Gene-1",
    "1st ranked Gene explanation": "<Gene-1 explanation>",
    "2nd ranked Gene": "Gene-2",
    "2nd ranked Gene explanation": "<Gene-2 explanation>",
    "3rd ranked Gene": "Gene-3",
    "3rd ranked Gene explanation": "<Gene-3 explanation>",
    "source": [
      "<source-1 URL>",
```

*"<source-2 URL>",*
*"<source-3 URL>"*
*]*
*}*
*]*


#### aiDIVA-meta

**Supplemental Table 6** All features used to train the aiDIVA-meta random forest model sorted according to their importance.

| Feature | Importance |
|---|---|
| eb_rank | 0.263248 |
| eb_rank_llm | 0.217983 |
| rf_rank_llm | 0.178370 |
| eb_score | 0.171753 |
| rf_rank | 0.082627 |
| rf_score | 0.078625 |
| recessive | 0.003699 |
| dominant | 0.003695 |

**Supplemental Table 7** Parameter settings used to train the aiDIVA-meta random forest model.

| Parameter | Value |
|---|---|
| bootstrap | True |
| ccp_alpha | 0.0 |
| class_weight | None |
| criterion | gini |
| max_depth | None |
| max_features | sqrt |
| max_leaf_nodes | None |
| max_samples | None |
| min_impurity_decrease | 0.0 |
| min_samples_leaf | 1 |
| min_samples_split | 2 |
| min_weight_fraction_leaf | 0.0 |
| n_estimators | 1000 |
| n_jobs | None |
| oob_score | False |
| random_state | 14038 |
| verbose | 0 |
| warm_start | False |

**Supplemental Table 8** All features used to train the aiDIVA-meta-RF random forest model sorted according to their importance.

| Feature | Importance |
|---|---|
| rf_rank_llm | 0.418360 |
| rf_score | 0.349883 |
| rf_rank | 0.231757 |

**Supplemental Table 9** Parameter settings used to train the aiDIVA-meta-RF random forest model.

| Parameter | Value |
|---|---|
| bootstrap | True |
| ccp_alpha | 0.0 |
| class_weight | None |
| criterion | gini |
| max_depth | None |
| max_features | sqrt |
| max_leaf_nodes | None |
| max_samples | None |
| min_impurity_decrease | 0.0 |
| min_samples_leaf | 1 |
| min_samples_split | 2 |
| min_weight_fraction_leaf | 0.0 |
| n_estimators | 1000 |
| n_jobs | None |
| oob_score | False |
| random_state | 14038 |
| verbose | 0 |
| warm_start | False |

### Overview of diseases in the benchmark set

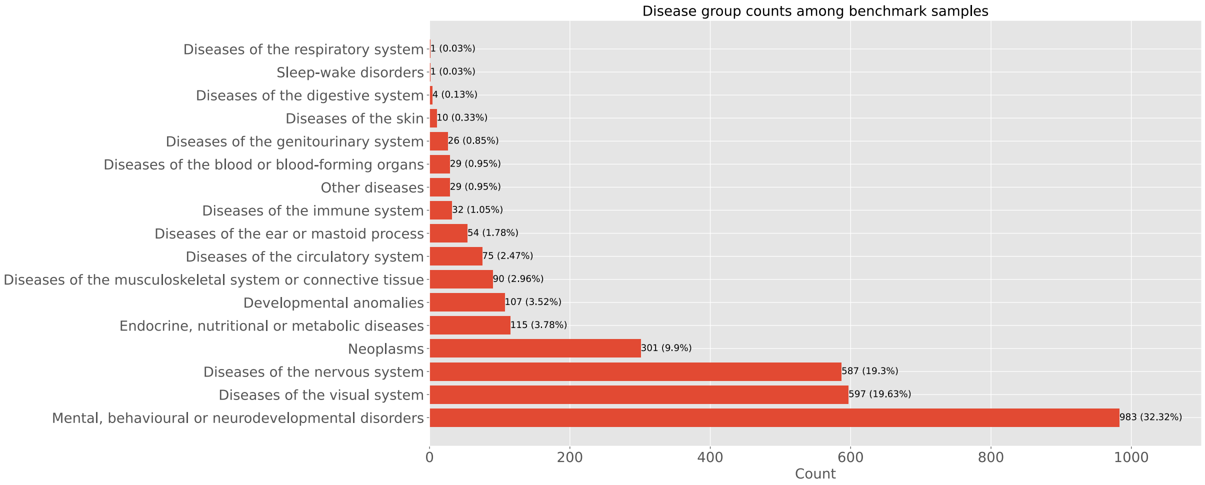


**Supplemental Figure 2** Overview of the disease groups present in our benchmark set (3041 cases).

#### References


1. McLaren W, Gil L, Hunt SE, et al. The Ensembl Variant Effect Predictor. *Genome Biol*. 2016;17(1):1-14. doi:10.1186/s13059-016-0974-4

2. Sturm M, Chernov A, Schütz L, et al. imgag/ngs-bits: 2025_03. Published online March 19, 2025. doi:10.5281/zenodo.15051584

3. Chen S, Francioli LC, Goodrich JK, et al. A genomic mutational constraint map using variation in 76,156 human genomes. *Nature*. 2024;625(7993):92-100. doi:10.1038/s41586-023-06045-0

4. Pollard KS, Hubisz MJ, Rosenbloom KR, Siepel A. Detection of nonneutral substitution rates on mammalian phylogenies. *Genome Res*. 2010;20(1):110-121. doi:10.1101/gr.097857.109

5. Siepel A, Bejerano G, Pedersen JS, et al. Evolutionarily conserved elements in vertebrate, insect, worm, and yeast genomes. *Genome Res*. 2005;15(8):1034-1050. doi:10.1101/gr.3715005

6. Ng PC, Henikoff S. Predicting Deleterious Amino Acid Substitutions. *Genome Res*. 2001;11(5):863-874. doi:10.1101/gr.176601

7. Adzhubei I, Jordan DM, Sunyaev SR. Predicting Functional Effect of Human Missense Mutations Using PolyPhen-2. *Curr Protoc Hum Genet*. 2013;76(1):7.20.1-7.20.41. doi:10.1002/0471142905.hg0720s76

8. González-Pérez A, López-Bigas N. Improving the Assessment of the Outcome of Nonsynonymous SNVs with a Consensus Deleteriousness Score, Condel. *Am J Hum Genet*. 2011;88(4):440-449. doi:10.1016/j.ajhg.2011.03.004

9. Rentzsch P, Witten D, Cooper GM, Shendure J, Kircher M. CADD: predicting the deleteriousness of variants throughout the human genome. *Nucleic Acids Res*. 2019;47(D1):D886-D894. doi:10.1093/nar/gky1016

10. Li S, van der Velde KJ, de Ridder D, et al. CAPICE: a computational method for Consequence-Agnostic Pathogenicity Interpretation of Clinical Exome variations. *Genome Med*. 2020;12(1):1-11. doi:10.1186/s13073-020-00775-w

11. Ioannidis NM, Rothstein JH, Pejaver V, et al. REVEL: An Ensemble Method for Predicting the Pathogenicity of Rare Missense Variants. *Am J Hum Genet*. 2016;99(4):877-885. doi:10.1016/j.ajhg.2016.08.016

12. Reva B, Antipin Y, Sander C. Predicting the functional impact of protein mutations: application to cancer genomics. *Nucleic Acids Res*. 2011;39(17):e118. doi:10.1093/nar/gkr407

13. Rogers MF, Shihab HA, Mort M, Cooper DN, Gaunt TR, Campbell C. FATHMM-XF: accurate prediction of pathogenic point mutations via extended features. *Bioinformatics*. 2018;34(3):511-513. doi:10.1093/bioinformatics/btx536

14. Cheng J, Novati G, Pan J, et al. Accurate proteome-wide missense variant effect prediction with AlphaMissense. *Science*. 2023;381(6664):eadg7492. doi:10.1126/science.adg7492

15. Bailey JA, Yavor AM, Massa HF, Trask BJ, Eichler EE. Segmental Duplications: Organization and Impact Within the Current Human Genome Project Assembly. *Genome Res*. 2001;11(6):1005-1017. doi:10.1101/gr.187101

16. Bailey JA, Gu Z, Clark RA, et al. Recent Segmental Duplications in the Human Genome. *Science*. 2002;297(5583):1003-1007. doi:10.1126/science.1072047

17. Benson G. Tandem repeats finder: a program to analyze DNA sequences. *Nucleic Acids Res*. 1999;27(2):573-580. doi:10.1093/nar/27.2.573

18. Nassar LR, Barber GP, Benet-Pagès A, et al. The UCSC Genome Browser database: 2023 update. *Nucleic Acids Res*. 2023;51(D1):D1188-D1195. doi:10.1093/nar/gkac1072

19. Smit A, Hubley R. RepeatMasker. Published online March 2025. https://github.com/Dfam-consortium/RepeatMasker

20. Karczewski KJ, Francioli LC, Tiao G, et al. The mutational constraint spectrum quantified from variation in 141,456 humans. *Nature*. 2020;581(7809):434-443. doi:10.1038/s41586-020-2308-7

21. Jaganathan K, Kyriazopoulou Panagiotopoulou S, McRae JF, et al. Predicting Splicing from Primary Sequence with Deep Learning. *Cell*. 2019;176(3):535-548.e24. doi:10.1016/j.cell.2018.12.015

22. Sturm M, Admard J, Schütz L, et al. imgag/megSAP: 2025_03. Published online March 21, 2025. doi:10.5281/zenodo.15063428

23. OpenAI, Hurst A, Lerer A, et al. GPT-4o System Card. *arXiv*. Preprint posted online October 25, 2024. doi:10.48550/arXiv.2410.21276
